# GLP-1 Receptor Agonists, Stroke, and Cognitive Outcomes: Systematic Review and Meta-Regression

**DOI:** 10.64898/2026.09.18.26363449

**Authors:** Marianna Danielli, Rhea Sibal, George Collett, Humza Awan, Cindy Sharma, James Steckelmacher, Cormac Kennedy, Ajay Gupta

## Abstract

**Importance:** Randomised-controlled trials (RCTs) suggest glucagon-like-peptide-1 receptor agonists (GLP-1RAs) reduce the risk of stroke in type 2 diabetes (T2DM); however, the mechanisms remain unclear. If benefits were driven principally by blood pressure (BP) lowering, similar effects on cognitive impairment may be expected. Evaluating data from trials with robust ascertainment and prespecified stroke and/or cognitive outcomes may provide greater clarity.

**Objective:** To confirm the effect of GLP-1RAs on stroke risk in RCTs with prespecified, adjudicated stroke endpoints; to explore whether risk reduction is associated with trial-level changes in systolic BP (SBP), glycated haemoglobin (HbA1c), or body weight; to establish whether a parallel cognitive signal exists.

**Data Sources:** PubMed, Cochrane, Embase, and CINAHL, from inception to August 2026 (PROSPERO CRD420261305938).

**Study Selection:** RCTs comparing GLP-1RAs with placebo/usual care in adults, with ≥12 months follow-up, and prespecified stroke and/or cognitive outcomes.

**Data Extraction and Synthesis:** Hazard ratios (HRs) for stroke were pooled using inverse-variance random-effect meta-analysis. Univariable meta-regression related logHR for stroke to trial-level between-group differences in SBP, HbA1c and weight, with permutation-based inference and leave-one-out sensitivity analyses. The pooled effect was benchmarked against that predicted from achieved SBP differences. Heterogeneous cognitive outcomes were synthesized narratively.

**Main Outcomes and Measures:** Adjudicated stroke events and prespecified cognitive endpoints.

**Results:** Among 7 RCTs (n=50,617), GLP-1RA therapy was associated with 17% lower stroke risk (pooled HR 0.83; [95% CI 0.75-0.92]; p<.001). Meta regression showed an association with SBP (β = 0.371; [95% CI 0.013-0.729]; p=.043), but not HbA1c (0.432; [-0.064-0.928]; p=.088) or weight (0.109; [-0.038-0.256]; p=.148). The mean between-group SBP difference (∼1.45 mmHg) predicts a stroke reduction of roughly 4%, versus the 17% observed, leaving approximately three quarters of the log-effect unexplained by BP lowering. Four trials (n=4,206) in established neurodegenerative disease showed no cognitive benefit.

**Conclusions and Relevance:** GLP-1RA therapy reduces stroke risk by 17% among individuals with T2DM at high cardiovascular risk, a benefit not fully accounted for by conventional cardiometabolic risk factor change. Additional mechanisms warrant evaluation. The absence of cognitive signal suggests the benefit may have cerebrovascular origin, but whether GLP-1RAs prevent vascular cognitive impairment in cardiometabolic populations remains untested.

## Introduction

Stroke and dementia are the leading global neurological causes of death and disability(1). Common, potentially modifiable cardiovascular risk factors - hypertension, obesity, type 2 diabetes (T2DM) - drive a large proportion of the stroke (90%) and dementia (45%) burden(2,3). T2DM increases stroke risk via microvascular injury and large-vessel atherosclerosis, and accelerates cognitive decline(4,5). Incident stroke is itself directly associated with accelerated cognitive decline and dementia, so stroke-preventing therapies may influence cognitive trajectories through that route alone(6).

Glucagon-like peptide-1 receptor agonists (GLP-1RAs) are widely used in T2DM and obesity for their glycemic and weight-lowering effects, and recommended for cardiovascular risk reduction in at-risk individuals(7). Cardiovascular outcome trials consistently show reductions in major adverse cardiovascular events, including stroke(8–10). Nonetheless, there are two key gaps.

First, existing meta-analyses typically pool trials without stroke as an adjudicated endpoint, risking ascertainment and outcome-reporting bias(10–12). Establishing the effect where stroke was prospectively defined and adjudicated is a necessary precondition for mechanistic inference.

Second, it is unclear what drives that benefit. GLP-1RAs modestly and consistently reduce systolic blood pressure (SBP) while also reducing glycated haemoglobin (HbA1c) and body weight. These changes could explain the stroke risk reduction, although it has rarely been examined(13). This can be tested by examining whether trials achieve larger cardiometabolic risk-factor reductions and whether the observed benefit is commensurate with that expected. Only one prior study investigated mediation through HbA1c and weight, finding no significant associations, but without assessing SBP(14). Preclinical and early clinical work also suggests direct anti-inflammatory, endothelial, and anti-amyloid actions operating beyond traditional stroke risk-factor modification(15,16).

These two candidate explanations (conventional cardiometabolic risk factor modification or neurobiological effects) render cognitive outcomes an informative and complementary endpoint. Reduced SBP is associated with cognitive impairment risk reduction, and cardiometabolic risk factor modification may therefore drive any benefits of GLP-1RAs on stroke and, in parallel, cognitive outcomes(17). However, if no cognitive signal emerges alongside a robust stroke benefit, this may suggest that GLP-1RAs preferentially prevent discrete cerebrovascular events rather than modifying the broader vascular and neurobiological processes underlying cognitive impairment. This inference is necessarily indirect: the interval between stroke and the emergence of cognitive impairment typically exceeds the duration of cardiovascular outcome trials, which are not designed to capture it. Cognitive outcomes therefore serve as a mechanistic probe into the pathways linking GLP-1RAs to cerebrovascular and cognitive health. To date, few studies have evaluated stroke and cognitive outcomes together, and to our knowledge none restricted to trials with prespecified, adjudicated endpoints while also examining whether any cerebrovascular benefit of GLP-1RAs is mediated through cardiometabolic risk-factor modification, or reflects direct neurobiological effects.

Accordingly, this systematic review has four aims: (1) to confirm the GLP-1RA therapy effect on stroke risk in randomised controlled trials (RCTs) with prespecified and adjudicated outcomes; (2) to examine whether stroke risk reduction is associated with trial-level changes in SBP, HbA1c, body weight; (3) to benchmark the observed benefit against that predicted from achieved cardio-metabolic risk-factor changes; and (4) to synthesise cognitive outcome from RCTs of GLP-1RAs with prespecified cognitive endpoints.

## Methods

This systematic review with meta-analysis was prospectively registered in PROSPERO (CRD420261305938), and followed PRISMA 2020 reporting guidelines. the completed PRISMA 2020 checklist is provided in eTable 1.

### Data Sources and Search Strategy

PubMed/MEDLINE, Cochrane, Embase, and CINAHL were searched for RCTs of GLP-1RAs, published from inception to August 2026 (eMethods). No language restrictions were applied.

### Study selection

RCTs were eligible if they met all of the following: (1) adult participants aged ≥ 18 years; (2) comparing GLP-1RA with placebo or usual care; (3) minimum follow-up duration of 12 months; (4) reporting pre-specified primary or key secondary trial endpoints of stroke (any adjudicated stroke event: ischemic, hemorrhagic, or unspecified), dementia (adjudicated or cognitive disorder diagnoses), or cognitive outcomes (assessed using validated cognitive assessments). Trials with an active comparator and trials reporting stroke only within a composite endpoint without a separately reported stroke-specific effect estimate, were excluded. On this basis, Harmony Outcomes (albiglutide) was excluded (eMethods). Trials were not excluded on the basis of population.

Two reviewers independently screened titles, abstracts and full texts, with disagreement resolved by a third reviewer.

### Data Extraction and Quality Assessment

Study characteristics were extracted from full texts and supplementary materials. Hazard ratios (HRs) and 95% confidence intervals (CIs) for stroke risk associated with GLP-1RAs vs. placebo/routine care were extracted from intention-to-treat analyses, or equivalent full analysis sets.

Trial-level between-group (GLP-1RAs vs. placebo/usual care) differences in the change in SBP, HbA1c and weight from baseline to post-intervention were extracted. Lipid fractions were extracted, but reported too inconsistently to support meta-regressions. Two reviewers extracted data independently, with discrepancies resolved by a third reviewer. Where cognitive outcomes lacked numerical reporting, corresponding authors were contacted.

Certainty of evidence was independently assessed using GRADE (Grading of Recommendations Assessment, Development and Evaluation) (eTable 2).

### Data Synthesis and Statistical Analysis

All analyses were conducted using R (version 4.6.1) using the metafor (version 5.0-1), readxl, dplyr, stringr, and readr packages(18).

For stroke outcomes, HRs were log-transformed and pooled with standard errors using inverse-variance methods. Random-effects meta-analyses used DerSimonian-Laird estimators for between-study variance. Fixed-effect models were run in parallel (eTable 3). Pooled effects were expressed as HRs with 95% CIs by exponentiating pooled logHRs.

Between-study heterogeneity was assessed using I² and τ², effect estimates in forest plots. Influence, leave-one-out funnel plots and Egger-type regression analyses were performed, recognising the limited number of trials.

Separate univariable trial-level random-effects meta-regression models were fitted with logHR for stroke as the dependent variable and ΔSBP (mmHg), ΔHbA1c (percentage points) and Δweight (kg) each entered as sole predictors. Covariates were modelled separately rather than jointly, since with 7 trials (6 for SBP) and strong collinearity between covariates (HbA1c-SBP r=0.92), a multivariable model would be over-parameterised. Because conventional inference is unreliable with few trials, we additionally reported exact permutation-based p-values and leave-one-out meta-regression. Fixed-effects meta-regression models are presented as supplementary analyses.

To assess whether the magnitude of the pooled effect was commensurate with the achieved BP difference, we compared pooled HRs for stroke risk predicted from the mean trial-level SBP difference with the association established by the Blood Pressure Lowering Treatment Trialists’ Collaboration (BPLTTC): a 5 mmHg SBP reduction corresponding to approximately 14% lower stroke risk(19). The proportion of the observed log-effect accounted for by SBP lowering was calculated as ratio of predicted to observed logHR. Because all included trials enrolled participants with T2DM, we used the estimate derived from participants with T2DM at baseline in the BPLTTC individual participant-level analysis, in effects were standardised to a 5-mmHg reduction in SBP using equivalent methods. This comparison was added post hoc and is indirect and illustrative rather than a formal mediation analysis.

Cognitive outcomes were synthesised narratively, as populations, cognitive assessment tools and outcome definitions differed markedly.

## Results

### Stroke Outcomes

Seven RCTs (N=50,617) were included (Figure 1), all in patients with T2DM at high cardiovascular risk. The GLP-1RAs included human-based analogues (e.g., liraglutide, semaglutide, dulaglutide) and exendin 4-based agents (e.g., exenatide, lixisenatide and efpeglenatide), with follow-up of 1.8 to 5.4 years(20–25). Baseline characteristics are shown in Table 1.

**Figure 1.**
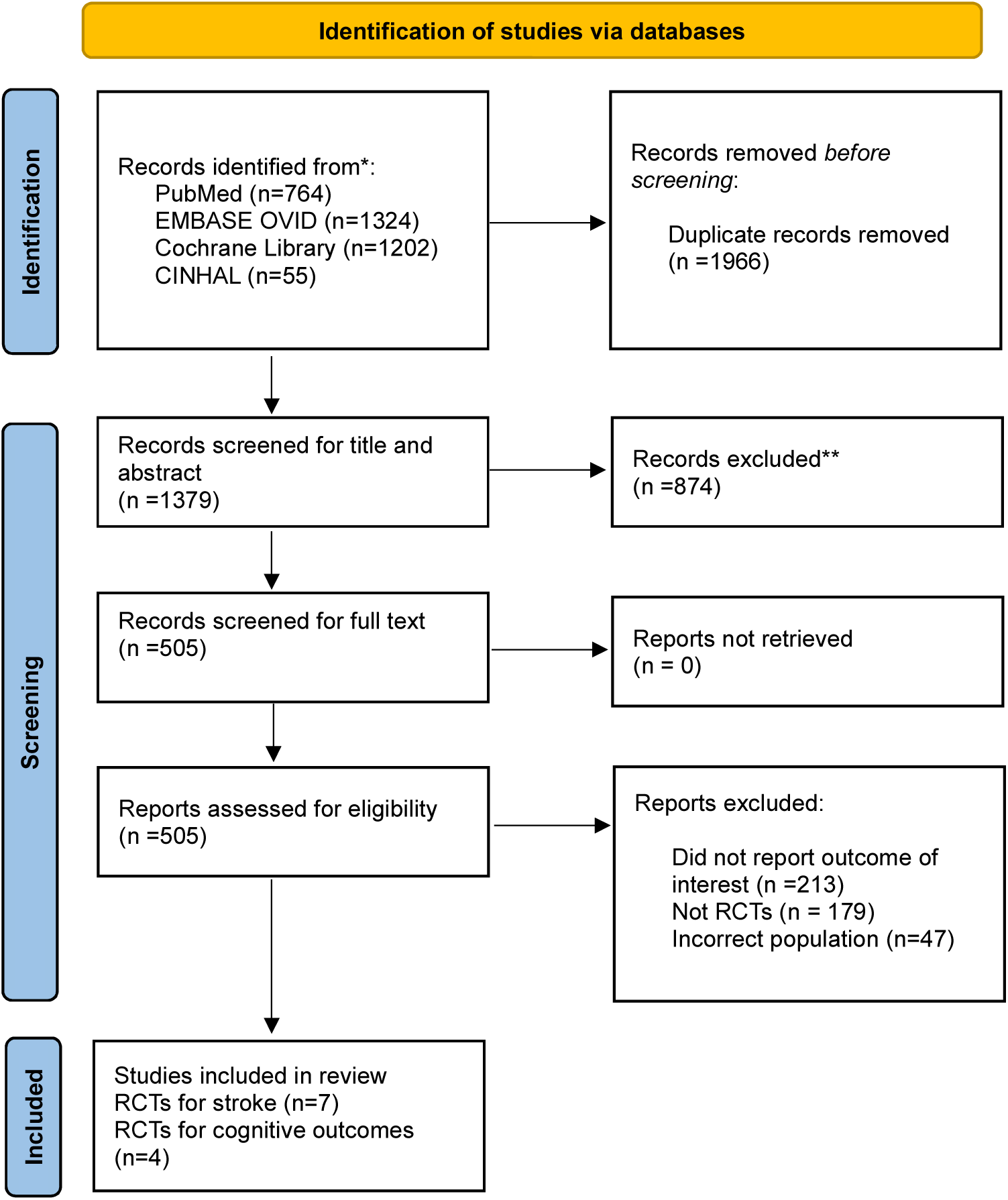
PRISMA Flowchart.

**Table 1.** Characteristics of Included GLP-1 Receptor Agonist Cardiovascular Outcome Trials.

| Source | No. of participants | Trial design | Study population | Mean age, (% of Female participants) | Intervention Group | Control Group | Median follow-up | Primary outcome (stroke prespecified as an adjudicated component) |
| --- | --- | --- | --- | --- | --- | --- | --- | --- |
| **ELIXA** (42) | 6,068 | Randomized, double-blind, placebo-controlled RCT | Adults with T2DM and recent acute coronary syndrome | 60.3, (30.7) | Lixisenatide once daily | Placebo | 25 months | 4-point MACE (CV death, non-fatal MI, non-fatal stroke, or hospitalization for unstable angina) |
| **LEADER**(20) | 9,340 | Randomized, double-blind, placebo-controlled RCT | Adults with T2DM at high cardiovascular risk | 64.3, (35.7) | Liraglutide up to 1.8 mg daily | Placebo |  | 3-point MACE (CV death, non-fatal MI, or non-fatal stroke) |
| **SUSTAIN-6** (24) | 3,297 | Randomized, double-blind, placebo-controlled RCT | Adults with T2DM at high cardiovascular risk. |  | Semaglutide 0.5 or 1.0 mg weekly | Placebo |  | 3-point MACE (CV death, non-fatal MI, or non-fatal stroke) |
| **EXSCEL**(43) | 14,752 | Randomized, double-blind, placebo-controlled RCT | Adults with T2DM with or without prior cardiovascular disease | 62, (38.1) | Exenatide ER 2 mg weekly | Placebo | 3.2 years | Time to first 3-point MACE (CV death, non-fatal MI, or non-fatal stroke) |
| **REWIND** (22) | 9,901 | Randomized, double-blind, placebo-controlled RCT | Adults with T2DM and either established CVD or multiple cardiovascular risk factors | 66.2, (46.4) | Dulaglutide 1.5 mg week | Placebo | 5.4 years | Time to first 3-point MACE (CV death, non-fatal MI, or non-fatal stroke) |
| **PIONEER 6** (44) | 3,183 | Randomized, double-blind, placebo-controlled RCT | Adults with T2DM at high cardiovascular risk | 66.0 (32.8) | Oral semaglutide 14 mg daily | Placebo | 19 months | Time to first 3-point MACE (CV death, non-fatal MI, or non-fatal stroke) |
| **AMPLITUDE-O** (25) | 4,076 | Randomized, double-blind, placebo-controlled RCT | Adults with T2DM and established cardiovascular disease or multiple risk factors | 64.5 (32.7) | Efpeglenatide 4 mg or 6 mg weekly | Placebo | 1.8 years | Time to first 3-point MACE (CV death, non-fatal MI, or non-fatal stroke) |

*MACE, major adverse cardiovascular events; CV, cardiovascular; MI, myocardial infarction; RCT, randomised controlled trial.*

There were 558 stroke events among 25,308 participants assigned to GLP-1RAs and 671 events among 25,309 participants assigned to placebo/usual care (Table 1).

GLP-1RA therapy was associated with a 17% stroke-risk reduction (pooled HR 0.83; 95% CI 0.75-0.92; p<.001), with no between-study heterogeneity (I²=0%; τ²=0) (Figure 2); the fixed-effect estimate was identical.

**Figure 2.**
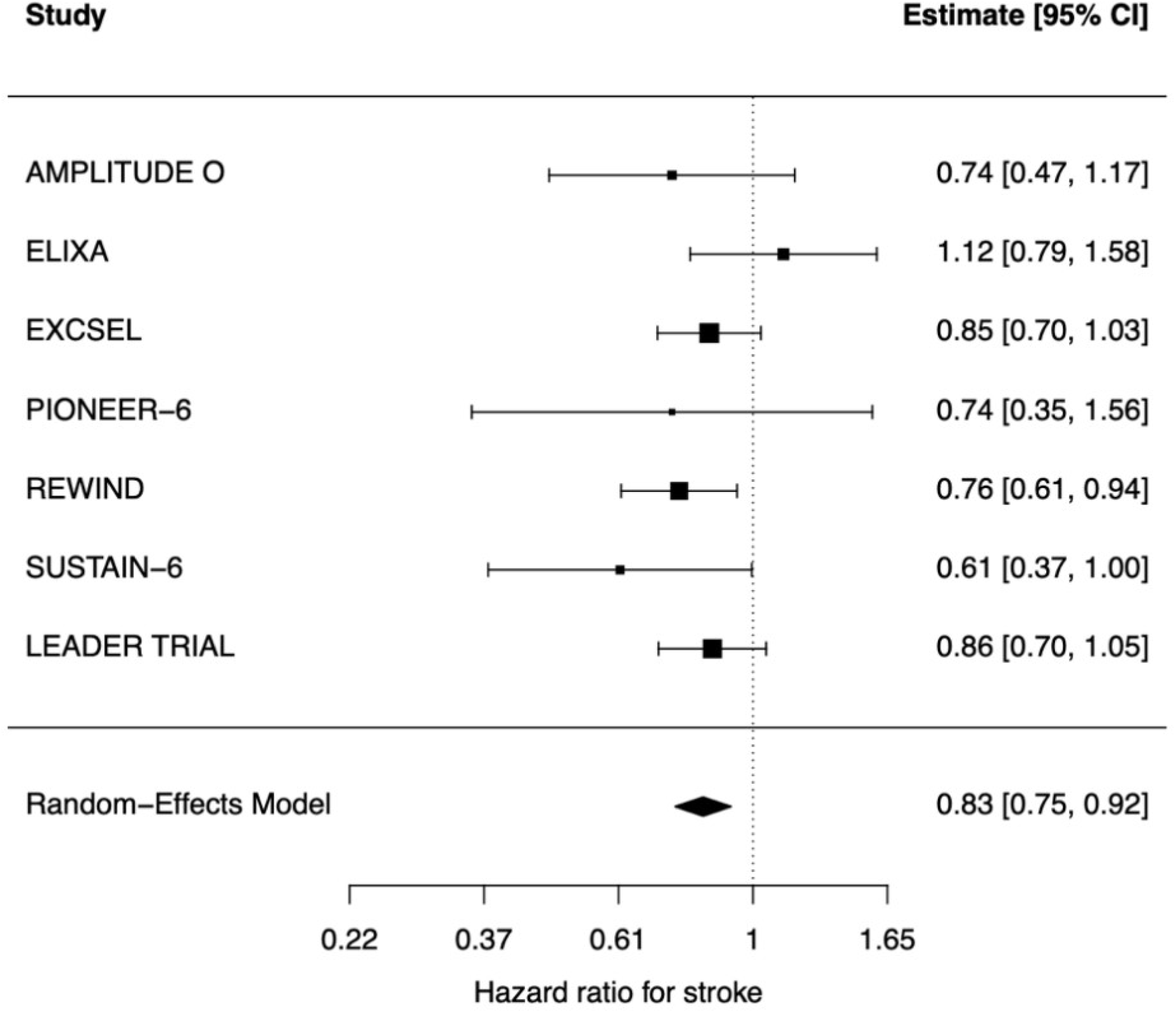
Total stroke data pooled result

Point estimates were concordant across six trials, ELIXA excepted (HR 1.12; [95% CI 0.79-1.58]). Estimates for ischaemic stroke, and for the composite of ischaemic stroke or transient ischaemic attack, where reported, were similar to the pooled effect; haemorrhagic stroke findings remained inconclusive.

Omission of any single trial did not materially alter the pooled estimate (range 0.81-0.85) with overlapping confidence intervals throughout (eFigure 1). Funnel plot inspection and Egger-type regression did not reveal clear small-study asymmetry evidence (eFigure 2).

This homogeneity was observed despite substantial between-trial variation in achieved cardiometabolic effect, with SBP differences ranging from approximately -0.80 to -1.95 mmHg, and correspondingly wide HbA1c and weight ranges (eTable 4).

### Determinants of Stroke Benefit

Trial-level meta-regression showed significant associations between stroke risk reduction and SBP changes (k=6; β=0.371 [95% CI 0.013 to 0.729]; p=.043), but not with HbA1c (k=7; p=.088) or weight (k=7; p=.148). Residual heterogeneity was zero in all three models (eTable 3, eFigures 3-5).

**Figure 3.**
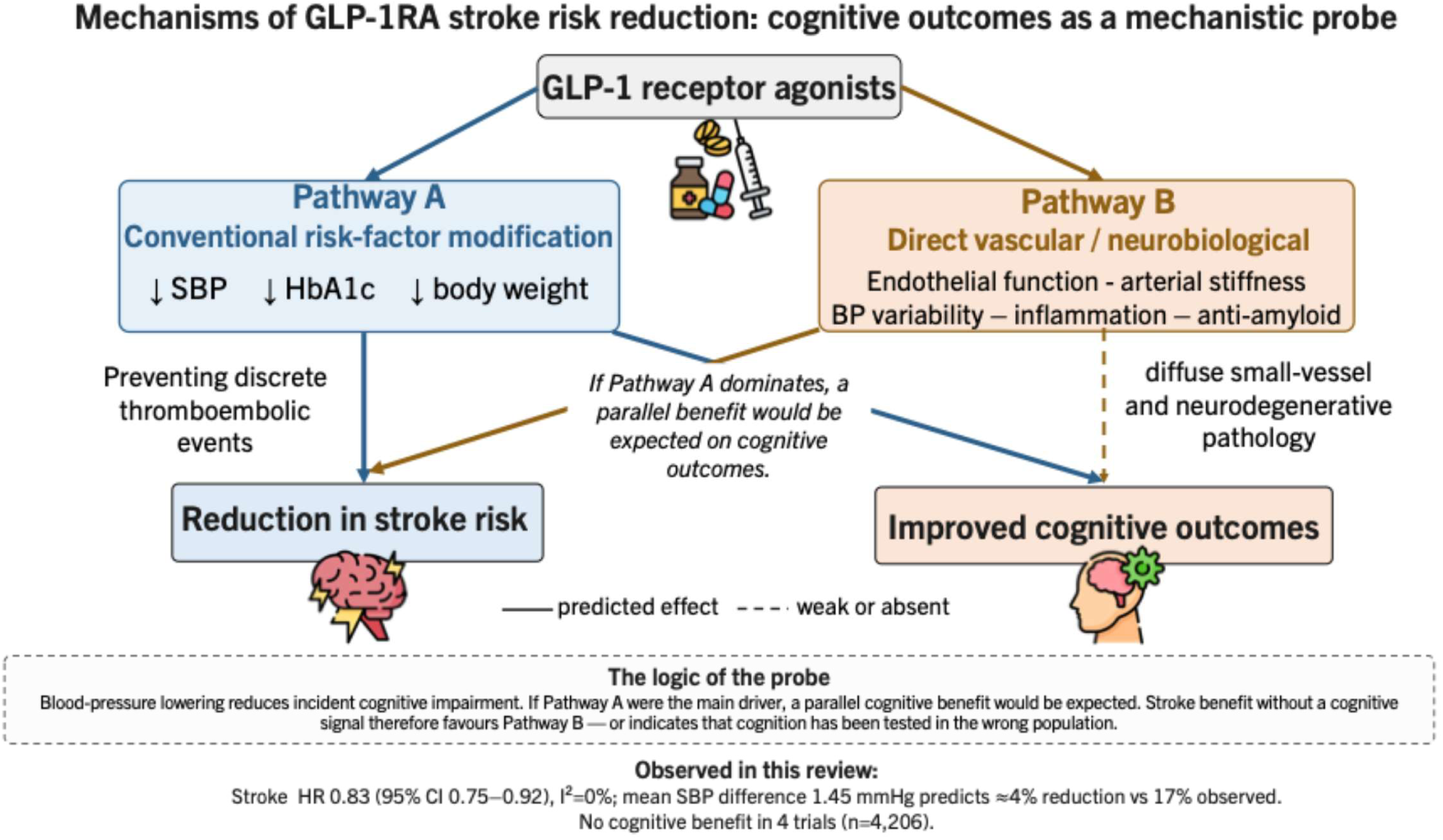
Conceptual model of candidate mechanisms linking GLP-1 receptor agonist therapy to cerebrovascular and cognitive outcomes. Pathway A operates through conventional cardiometabolic risk-factor modification; Pathway B through direct vascular and neurobiological actions. Because BP-lowering reduces incident cognitive impairment, a benefit operating principally through Pathway A would be expected to produce parallel effects on both endpoints. A stroke benefit without a corresponding cognitive signal is therefore consistent with Pathway B, but cannot confirm it, since all cognitive trials to date enrolled participants with established neurodegenerative disease. The four cognitive trials were substantially heterogeneous in population and underlying neuropathology (Alzheimer’s disease in three and Parkinson’s disease in one) and none enrolled participants with vascular cognitive impairment.

Sensitivity analyses showed that the SBP association was retained on exact permutation testing (p=.017; eTable 5), but it was not robust to the exclusion of individual trials: on leave-one-out meta-regression, the association was non-significant when ELIXA (p=.255), SUSTAIN-6 (p=.103), and REWIND (p=.076) were omitted (eTable 6).

Baseline mean SBP was examined on the reasoning that trials enrolling participants with higher baseline BP might achieve larger reductions and hence larger benefit. It was not significantly associated with stroke risk reduction (p=.068), although the direction was consistent with greater benefit at higher baseline pressure, and neither term remained significant when entered together (omnibus p=.107), reflecting collinearity (r=-0.78). Baseline SBP spanned only 129.5 to 137.0 mmHg across trials (eTable 7).

Benchmarking the pooled effect against achieved BP differences indicated a substantial shortfall. The mean trial-level between-group SBP difference was approximately 1.45 mmHg, which, applying the BPLTTC estimate of 14% stroke reduction per 5 mmHg among participants with T2DM, predicts stroke risk reduction of approximately 4% (predicted HR 0.96)(19). This assumes the log-linear SBP-stroke relationship demonstrated in large prospective cohort and trial meta-analyses, which holds to at least 115 mmHg without a threshold(26,27). The observed reduction was 17% (HR 0.83). SBP lowering therefore accounts for approximately one quarter of the observed log-effect, leaving approximately three quarters unexplained. Even taking the largest single-trial SBP difference (1.95 mmHg), the predicted reduction reaches only approximately 6%.

### Cognitive Outcomes

Four trials (N=4,206) reporting prespecified cognitive endpoints were identified (Table 2). None demonstrated cognitive benefit.

**Table 2.** Characteristics of Randomised Trials Reporting Prespecified Cognitive Outcomes of GLP-1 Receptor Agonists.

| Source | No. of participants | Trial design | Study population | Mean age, (% of Female participants) | Intervention Group | Control Group | Follow-up in months | Primary outcome and cognitive endpoint |
| --- | --- | --- | --- | --- | --- | --- | --- | --- |
| **evoke(28)** | 1,855 | Randomized, double-blind, placebo-controlled phase 3 RCT | Adults 55-85 years old with MCI or mild dementia due to Alzheimer's disease, amyloid-positive | 71.8 (53.0) | Oral semaglutide 14 mg (flexible dose) once daily | Placebo | 24 | Change in Clinical Dementia Rating - Sum of Boxes (CDR-SB) from baseline to week 104. Result: estimated difference -0.08 (95% CI - 0.35 to 0.20); p=.57. |
| **evoke+(28)** | 1,953 | Randomized, double-blind, placebo-controlled phase 3 RCT | As evoke, additionally including participants with significant small-vessel pathology | 72.6 (51.9) | Oral semaglutide 14 mg (flexible dose) once daily | Placebo | 24 | Change in CDR-SB from baseline to week 104. Result: estimated difference 0.10 (95% CI -0.17 to 0.38); p=.46. |
| **ELAD(45)** | 204 | Randomized, double-blind, placebo-controlled RCT | Adults with mild-moderate Alzheimer's disease | 69.9 (52) | Liraglutide 1.8 mg once daily | Placebo | 12 | Primary: change in cerebral glucose metabolic rate (FDG-PET). Result: difference -0.17 (95% CI -0.39 to 0.06); p=.14. Secondary cognitive: ADAS-Exec +0.15 (0.03 to 0.28), unadjusted p=.01; CDR-SB -0.06 (-0.57 to 0.44), p=.81; ADCS-ADL -0.58 (-3.13 to 1.97), p=.65. |
| **Exenatide Parkinson's trial (30)** | 194 | Randomized, double-blind, placebo-controlled RCT | Adults with Parkinson's disease | 60.6 (38) | Exenatide 2 mg once weekly | Placebo | 24 | Primary: MDS-UPDRS Part III motor score in the off-medication state (primary endpoint not met). Secondary cognitive: Montreal Cognitive Assessment (MoCA) showed no between-group difference. |

*MCI = mild cognitive impairment; CDR-SB = Clinical Dementia Rating-Sum of Boxes; ADAS-Exec = Alzheimer's Disease Assessment Scale-Executive domain; ADCS-ADL = Alzheimer's Disease Cooperative Study-Activities of Daily Living; FDG-PET = fluorodeoxyglucose positron emission tomography; MDS-UPDRS = Movement Disorder Society Unified Parkinson's Disease Rating Scale.*

The evoke and evoke+ trials randomised 3,808 participants with early-stage symptomatic, amyloid-positive Alzheimer’s disease to oral semaglutide 14 mg or placebo for up to 156 weeks(28). Neither met primary endpoints: estimated between-group differences in the change in Clinical Dementia Rating–Sum of Boxes from baseline to week 104 were -0.08 (95% CI -0.35 to 0.20; p=.57) in evoke and 0.10 (-0.17 to 0.38; p=.46) in evoke+. Notably, evoke+ enrolled participants with significant small-vessel pathology and also showed no benefit.

In ELAD, liraglutide 1.8 mg daily versus placebo over 52 weeks in 204 participants with mild-to-moderate Alzheimer’s disease did not meet its primary endpoint of change in cerebral glucose metabolic rate (difference -0.17; 95% CI -0.39 to 0.06; p=.14)(29). A secondary executive-function measure (ADAS-Exec) favoured liraglutide (0.15; 0.03 to 0.28; unadjusted p=.01), but the trial was not powered for cognition, the p value was unadjusted for multiplicity, and neither Clinical Dementia Rating-Sum of Boxes (-0.06; -0.57 to 0.44; p=.81) nor Alzheimer’s Disease Cooperative Study-Activities of Daily Living differed between groups(29). In the Parkinson’s disease trial, once-weekly exenatide 2 mg over 96 weeks showed no significant between-group differences in Montreal Cognitive Assessment score or any other secondary cognitive or non-motor outcome(30).

## Discussion

In 7 RCTs (N=50,617) with prespecified, adjudicated endpoints, GLP-1RA therapy reduced stroke risk by 17%, a benefit not fully accounted for by accompanying changes in SBP, HbA1c or weight. Two evidence gaps follow: additional mechanisms underlying cerebrovascular GLP-1RAs benefits, and whether the benefit extends to preserving cognition in high cardio-metabolic risk individuals. Our estimate is consistent with recent cohort studies and meta-analyses, but was obtained under stricter inclusion criteria and is less vulnerable to ascertainment and outcome-reporting bias(12,31,32).

Conventional GLP-1RA risk-factor modifications do not appear to fully account for the stroke benefit. First, only SBP was associated with the magnitude of stroke risk reduction, which was retained on exact permutation testing but not on leave-one-out testing; HbA1c and weight showed no significant association. Second, the observed 17% reduction is more than four times larger than the approximately 4% predicted from the 1.45 mmHg mean SBP difference achieved. Third, the between-trial heterogeneity absence provides limited support for a simple dose-response model: trials achieving SBP differences varying more than two-fold produced similar stroke effects. Overall, SBP only partly accounts for the cerebrovascular benefit (Figure 3). Future trials should report additional vascular parameters.

Several alternative mechanisms merit consideration. GLP-1RAs improve endothelial function, arterial stiffness and may reduce BP variability, a determinant of stroke risk largely independent of mean BP(33). Lipid effects were reported too inconsistently across trials to permit meta-regression: future trials should report lipid fractions in a standardised manner. The absence of an association with glycaemic control is consistent with the broader literature: a meta-analysis of nine RCTs (n=59,197) found no effect of intensive glucose lowering on incident stroke (RR 0.96; 95% CI 0.88-1.06), and in the CONTROL individual participant-data analysis the 9% reduction in major cardiovascular events was driven by myocardial infarction rather than stroke(34,35). An earlier meta-regression reported such an association, which we did not reproduce(36). It differed from ours in three respects: it pooled 18 cardiovascular outcome trials spanning three drug classes (DPP-4 inhibitor, GLP-1RA and SGLT-2 inhibitor trials), so any HbA1c-stroke association cannot be attributed to GLP-1RAs specifically and is confounded by drug class; it used non-fatal stroke as a MACE component rather than requiring stroke to be prespecified and separately adjudicated; it examined glycaemia alone, without assessing BP or weight, so relative contributions could not be established. Its reported R² of 100% across a small number of trials is also consistent with overfitting (36).

The cognitive outcomes must be interpreted cautiously. No signal of cognitive benefit was observed across the four trials, all of which enrolled participants with established neurodegenerative diseases, in whom the underlying neuropathology may have been too advanced for any neuromodulatory benefit to emerge within the trial duration. The four trials also differed fundamentally in underlying neuropathology: evoke, evoke+ and ELAD enrolled participants with Alzheimer’s disease, in which β-amyloid and tau pathology predominate, while the exenatide trial enrolled participants with Parkinson’s disease, underpinned by α-synuclein pathology (28,30). None targeted vascular cognitive impairment, in which cerebral small-vessel disease is the principal substrate, accounting for approximately 20% of dementia cases. Cognitive impairment is therefore not a single entity, and the populations studied are not those in whom a cerebrovascular mechanism would manifest most readily. A more relevant test would be whether GLP-1RAs reduce incident vascular cognitive impairment in cardiometabolic populations followed over sufficient time, rather than whether they modify established protein-misfolding diseases. Taken together, these findings provide little evidence of direct cognitive benefit in established neurodegenerative diseases, while remaining compatible with a predominantly cerebrovascular mediated mechanism in the population where it would be expected to operate.

This aligns with Koychev and Holman, who distinguish treatment of symptomatic disease from prevention of incident dementia; once the clinical phase of Alzheimer’s disease is reached, substantial neuronal loss has occurred, limiting the capacity of any intervention to alter trajectories, so failure to show symptomatic benefit does not invalidate preventive mechanisms operating earlier in the disease course(37). They note that the established reduction with GLP-1RAs provides a plausible cerebrovascular pathway for modifying dementia risk, and that exenatide reduced circulating inflammatory proteins linked to Alzheimer’s disease pathophysiology in a post hoc analysis of EXSCEL, supporting immunomodulatory mechanisms(37,38).

Our finding that stroke benefits exceed what conventional risk-factor changes explain lends quantitative support to that cerebrovascular hypothesis, which is now being tested directly in the Impact of Semaglutide in Amyloid Positivity (ISAP) trial in cognitively unimpaired, amyloid-positive participants (39). Whether GLP-1RAs reduce the risk of vascular cognitive impairment or dementia in populations with T2DM, obesity, and elevated cardiovascular risk remains uncertain and warrants evaluation in appropriately designed trials with prespecified cognitive endpoints and prolonged follow-up. Observational evidence continues to suggest lower dementia risk with GLP-1RA exposure, highlighting discrepancies between trial and real-world data(40).

## Strengths and limitations

We restricted inclusion to trials with prespecified, adjudicated stroke endpoints, reducing ascertainment bias; risk of bias was low, and certainty of evidence high. We also interrogated mechanism, combining meta-regression, external benchmarking, and the cognitive comparison.

Several limitations warrant consideration. First, trial-level rather than individual-level data limited power and precluded formal mediation analysis, and the small number of trials necessitated separate univariable models. Second, zero between-study heterogeneity and modest SBP, HbA1c and weight variation limited the variance available to explain, may render coefficients fragile and, with unmeasured trial-level confounding, constrain inferences about mediators; we therefore de-emphasised the nominal SBP association and reported permutation-based inference. BP variability, endothelial function and lipid fractions were not reported in a form permitting trial-level analysis, so their role remains inferred. Third, the benchmarking comparison assumes that a given reduction in SBP confers the same stroke risk reduction, whether achieved with GLP-1RAs or with antihypertensive agents, and the relationship is linear. Fourth, the modest between-group SBP differences should not be read as the maximum attainable effect: participants were largely normotensive, and larger reductions might be expected in hypertensive populations, who are also at greatest stroke risk(13). Moreover, background antihypertensive therapy is adjusted in both arms of closely monitored trials, attenuating the observed between-arm difference and biasing our benchmarking towards underestimating benefits beyond BP lowering(13). Finally, all cognitive trials were conducted in neurodegenerative rather than cardiometabolic populations, so the absence of a cognitive signal cannot exclude benefit in the population in which the stroke benefit was observed. Given that statins have been associated with reduced dementia risk, this represents a clinically important line of inquiry(31,41).

## Conclusion

In trials with prespecified adjudicated endpoints, GLP-1RA therapy reduced stroke risk by 17% among T2DM, high cardiovascular risk individuals. This benefit may not be fully accounted for by accompanying reductions in SBP, HbA1c or body weight: SBP lowering explained approximately one quarter of the observed effect, whereas glycaemic and weight changes show no trial-level association. Additional mechanisms including improved endothelial function, reduced BP variability, and anti-inflammatory vascular effects may contribute and warrant further evaluation. We found no parallel cognitive benefit across four trials of populations with established neurodegenerative disease. Whether GLP-1RAs prevent vascular cognitive impairment in cardiometabolic populations remains untested and should be the priority for future trials with prespecified cognitive endpoints and long-term follow-up.

## Data Availability

All available data are in the Manuscript and Supplemental Data File. Statistical analysis code will be made available upon request.

## Acknowledgements

A.K.G. developed the study concept. R.S., H.A., and C.S. conducted the electronic search, screening, study selection and data extraction. M.D. conducted statistical analyses, interpreted results and wrote the majority of the manuscript. G.C. contributed to statistical analyses and paper writing-up. J.S. contributed to electronic search and paper writing-up. All the authors revised, commented on, read and approved the final version of the manuscript.

## Sources of Fundings

This work was performed as part of a Biomedical Research Centre (BRC) and National Institute for Health and Care Research (NIHR) funded programme. MD acknowledges the support of the NIHR Academic Clinical Fellowship (NIHR ACF-2024-19-010), the Barts Charity ACF Support Grant (G-003218), Barts Health NHS Trust, and Queen Mary University of London. AKG is supported by the National Institute for Health and Care Research (NIHR202116) and the NIHR Barts Biomedical Research Centre (NIHR203330). The funders had no role in the design, conduct, analysis, interpretation, or reporting of the study. The views expressed are those of the authors and not necessarily those of the NIHR or the Department of Health and Social Care.

## Disclosures

The authors have nothing to disclose.

## Notes

### Competing Interest Statement

The authors have declared no competing interest.

### Clinical Trial

n/a

